# Exploring sex-specific causal links between thousands of proteins and lipid metabolism using the UK Biobank Pharma proteomics data

**DOI:** 10.1101/2025.09.16.25335948

**Authors:** D. Zanetti, M. Koprulu, F. Grosso, D.V. Zhernakova, F. Crobu, S. Sanna

## Abstract

Lipid traits are known to be sex-differential, but the underlying molecular players are largely unknown. Since protein levels are downstream products of gene expression, proteomics data can be crucial to understand etiology of sex-differences in lipids metabolism.

We used sex-specific pQTL summary statistics for 2,923 circulating proteins measured in the UK Biobank Pharma Proteomics Project, along with sex-specific summary statistics of lipids from the Global Lipids Genetics Consortium. We combined these using two-sample Mendelian Randomization analyses and applied stringent multiple testing p-values correction and sensitivity analyses. We identified several sex-specific significant causal links between protein levels and lipid phenotypes: 82 were exclusive to females and 82 to males. The estimated causal effect was instead significant in both sexes but substantially different in size for 38 causal relationships (p <0.05). In the Lifelines Cohort, we replicated 16 sex-specific causal relationships and 4 sex-differentiated relationships also including GWASs restricted to non-statin users, ruling out the potential confounding of lipid-lowering medications more often prescribed in males. Intriguingly, several of these proteins were previously shown to be involved in inflammation and cardiometabolic disease, such as Apolipoprotein(a) and lipoprotein lipase. These results provide insight into potential genetic drivers of sex dimorphism in lipid metabolism.

## Introduction

Cardiovascular diseases (CVDs) are still the primary cause of death worldwide, with complex diagnosis and treatment further complicated by large sex differences in age of onset, symptoms and disease course ^1–3^. Response to treatment can also vary drastically between males and females, mostly because many current therapies are based on clinical and pre-clinical research studies largely based on male subjects ^2^. The existing knowledge gap of sex-specific or sex-differential biological mechanisms hampers the application of personalized medicine, leaving females underdiagnosed and undertreated.

One of the most important biomarkers of CVD is circulating lipid levels, for which epidemiological studies have highlighted substantial differences between sexes ^4^. It is well known, for example, that pre-menopausal females have lower levels of low-density lipoprotein cholesterol (LDL) than same age males ^5^ however, after menopause, this pattern tends to reverse, with females having higher levels than males. In addition, high-density lipoprotein cholesterol (HDL) levels are higher among females of all ages compared to males ^5^. Lipid levels also show a greater estimated heritability in females compared with males ^6^ especially for LDL and total cholesterol (TC) (> 1.3-fold difference), underling a possible genetic component behind this difference. Understanding the genetic determinants of sex difference in lipid levels and their molecular derivates could provide mechanistic insights into the biological processes underpinning CVD in both males and females and may improve CVD risk assessment and treatment.

Large scale genome-wide association studies conducted in millions of individuals, such as those carried out within the Global Lipid Genetic Consortium (GLGC) have highlighted the existence of genetic variants that are differentially associated with lipid levels between males and females ^7^. However, their molecular derivates and if and how these impact lipid levels in each sex, remain poorly understood.

Herein, we aim to highlight differential molecular signatures in lipid metabolism between males and females by identifying proteins that are causally linked to lipids in a sex-specific or sex-differentiated manner. Since proteins are downstream products of gene expression and can be more directly related to biological processes, proteomics can be crucial to understand sex-differences in lipids metabolism and thus ultimately pinpoint to similar mechanisms in CVD.

To this end, we used publicly available sex-stratified summary statistics of genetic determinants of protein and lipid levels, coupled with causal inference analyses. Specifically, we used sex-stratified cis-acting protein quantitative trait loci (pQTLs) summary statistics based on the UK Biobank Pharma Proteomics Project (UKB-PP) ^8^ proteomic measurements to infer causal relationships between a total of 2,923 circulating proteins and five lipid levels using the sex stratified GWAS summary statistics from GLGC ^7^. In addition, we used data from the Lifelines cohort to assess replication of our findings and to rule out the potential impact introduced by the known higher use of statins in males compared to females ^9,10^.

We highlighted several proteins that exhibited significant sex-differentiated causal relationships with lipids, mostly involved in pathways related with CVD, insulin and inflammation. The discovery of these sex-differences provides important etiological insights into the regulation of lipid levels. Such knowledge may, in turn, demonstrate the importance of sex-specific pQTLs and point to new or revised therapeutic sex specific strategies to prevent CVD.

## Materials and Methods

### Populations studied

In this study we used the sex-specific cis-pQTL GWAS summary statistics of 2,923 circulating proteins measured in the UKB-PPP, a precompetitive consortium of 13 biopharmaceutical companies ^11^, as exposure for our causal inference analyses ^8^. The UK Biobank proteomic measurements were conducted by antibody-based Olink technology Explore 1536 platform which uses Proximity Extension Assay ^12^. Further details about antibody-based proteomic measurements and quality control have been described elsewhere ^11^. Further details about the sex-stratified proteomic GWASs have also been described elsewhere ^8^. Briefly, proteins were available for 48,017 individuals (25,904 females and 22,113 males, aged 49–60). Protein abundances were inverse rank normalized and adjusted for technical covariates (e.g., blood draw timing, fasting status). Sex-stratified proteomic GWASs were then conducted using these residuals using REGENIE v.3.4.153 including age, age², and proteomic batch as covariates ^8^. Only individuals with matching sex chromosome and recorded/self-reported sex were included to avoid potential genotyping issues ^8^.

We also used sex-specific GWAS summary statistics of HDL, non-high-density lipoprotein cholesterol (non-HDL), LDL, total cholesterol (TC) and TG, downloaded from the GLGC (https://csg.sph.umich.edu/willer/public/glgc-lipids2021/results/sex_and_ancestry_specific_summary_stats/), ^7^ as outcomes. Sample size for these five traits varied between 459,578 and 581,618 for females and between 401,908 and 713,344 for males. In addition, we used the Lifelines cohort, a large biobank from Northern Netherlands ^9,13^, to replicate the significant findings using the same pipeline as implemented in the GLGC to derive GWAS statistics for lipids. Furthermore, we used this cohort to evaluate the impact of statins (lipid-lowering medications) and derived additional GWAS statistics after excluding individuals under such medications.

### GWAS of lipid levels in the Lifelines cohort

Lifelines Cohort Study data was used for replication of our main results and to evaluate the potential impact of statin usage in the results. Lifelines Cohort Study is a multidisciplinary prospective population-based cohort study that utilizes a unique three-generation design to investigate health and health-related behaviors in 167,729 individuals from the northern Netherlands. Lifelines employs a broad range of investigative procedures to assess the biomedical, socio-demographic, behavioral, physical, and psychological factors that contribute to health and disease, with a special focus on multi-morbidity and complex genetics ^9,13^.

Overall, 64,197 Lifelines individuals had both phenotype and genotype data available. We conducted sex-stratified GWAS in 2 groups: all individuals, regardless of the medication use, and individuals who did not report statin usage (as determined by ATC code C10AA) (N = 60,875). Percentage of females was 60% and 61 % in the two groups respectively.

Lipid levels data were processed in accordance with Graham et al. ^7^: 1) we log-transformed triglyceride levels; 2) we adjusted each lipid in each phenotype group and each sex for covariates by regressing out age, age squared and taking the residuals of the linear model; 3) we inverse-rank transformed these residuals. For the analysis of all individuals together irrespective of their medication status, we adjusted the lipid levels of individuals on statins to estimate their pre-medication levels by dividing the LDL cholesterol value by 0.7 and the total cholesterol value by 0.8.

Genotype data of Lifelines participants were generated in two batches. Batch one was genotyped on the Infinium Global Screening Array MultiEthnic Diseases version 1.0 and imputed using HRC v 1.1 as the reference. Detailed data processing description can be found in Lopera-Maya et al. ^14^. The second batch of samples was genotyped on the FinnGen Thermo Fisher Axiom® custom array and imputed using HRC v 1.1 as the reference. These two genotype batches were merged and filtered keeping genetic variants with minor allele frequency (MAF) > 0.05, call rate > 0.95, Hardy-Weinberg equilibrium p-value > 1e-6.

GWAS was done using REGENIE v3.4.1 ^15^: the first step (whole genome regression) was run on a subset of SNPs genotyped on the GSA array, with parameters--bsize 200--loocv; the second step was run on all imputed filtered SNPs using--bsize 200 parameter. Genotyping batch was used as covariate in both steps.

### Mendelian randomization analyses

We performed linkage disequilibrium (LD) clumping using a 10 Mb window and an r^2^ cutoff <= 0.001 for all the proteins-related GWAS available with UKB-PPP proteomic measurements, to select common and genome-wide significant independent hits; this step was carried out with PLINK (version 2.0) ^16^.

The variants selected by PLINK were then used as instrument variables (IVs) to perform two-sample Mendelian randomization (MR) analyses ^17^. MR was carried out using only in cis-acting variants. Cis pQTLs were defined as genetic variants located within 500 kb of the gene encoding the protein. We focused only on proteins encoded by genes located on autosomal chromosomes since MR on sex chromosomes would be difficult to interpretate given the inactivation of X chromosome. We performed the main MR analyses using the combined (all samples) as well as the sex-stratified pQTL GWAS summary statistics from the UKB-PPP as exposures ^8^ and the combined as well as the sex-stratified GWAS summary statistics of HDL, non-HDL, LDL, TC and TG from the GLGC ^7^ as outcomes. When IVs were absent in the outcome dataset, we utilized the LDproxy function in R ^18^ to identify suitable proxies in linkage disequilibrium (r2>0.8).

We performed two-sample MR using the standard inverse-variance weighted (IVW) regression - or the Wald ratio method in case of a single instrument variable - as main methods to declare significance. In addition, when multiple IVs were available, we used MR-PRESSO ^19^ to minimize the risk of horizontal pleiotropy, and the weighted median-based method. We specifically tested for presence of horizontal pleiotropy in all IVs by calculating intercept values from the MR Egger regression ^20^. Furthermore, we visually inspected scatter plots across all MR methods tested in our study and performed leave-one-out sensitivity analyses to identify if a single SNP was driving an association. We performed the two-sample MR analyses with the R package TwoSampleMR, version 0.6.8 ^17^.

We defined a causal relationship significant only if the following conditions were met: 1) p-value ≤ 0.05 after false discovery rate (FDR) correction (Benjamini-Hochberg method) ^21^ in IVW or Wald ratio, 2) nominal p-value < 0.05 for sensitivity MR methods MR-PRESSO and Weighted Median, and 3) MR-Egger intercept p>0.05 and 4) the association was not driven by a single SNP as determined by leave-one-out analysis and this SNP had not been removed by MR-PRESSO.

Since for MR conducted on one single IV sensitivity analyses cannot be performed, we further screened the Wald ratio results (the results of MR conducted on single IVs) and kept only proteins explaining at least 1% of the total variance in protein levels. Among the significant causal relationships detected, we defined “sex-specific” those relationships that met the significance criteria above (all four conditions) in one sex and the unadjusted p-value in main methods (IVW or Wald ratio) was ≥0.05 in the other sex.

As additional sensitivity analysis for the selected causal associations, we performed reverse MR to confirm the lack of significant reverse causation or confounding. To this end, we used the sex-stratified GWAS of lipids as exposure and sex-stratified GWAS of protein levels as outcome and selected IVs applying the same pruning criteria described above.

Since many protein-lipid relationships may be significant in one sex but still show nominal significance in the other sex or be associated with both sexes in a differentiated manner, we systematically evaluated differences in causal estimates detected in males and females. Specifically, we applied the Cochran’s Q test ^22^ to test for significant differences in the causal estimates from IVW (or Wald ratio for single variants) and selected those showing Cochran Q p-value < 0.05.

Finally, we performed replication of significant findings and evaluation of the influence of statin therapy by repeating the MR analyses using the same exposures (proteins), and as outcomes sex-stratified GWAS summary statistics of HDL, non-HDL, LDL, TC and TG derived from the Lifelines cohort. We used the following criteria to define replication: IVW, Wald ratio, Weighted median, MR-PRESSO p value ≤ 0.05, all leave-one-out p<0.05 and MR Egger intercept p value> 0.05 in the same sex-specific analysis as in the main analysis, and same direction of causal effect.

A flow chart of the different data sources and methods used in this study is shown in Figure 1.

**Figure 1.**
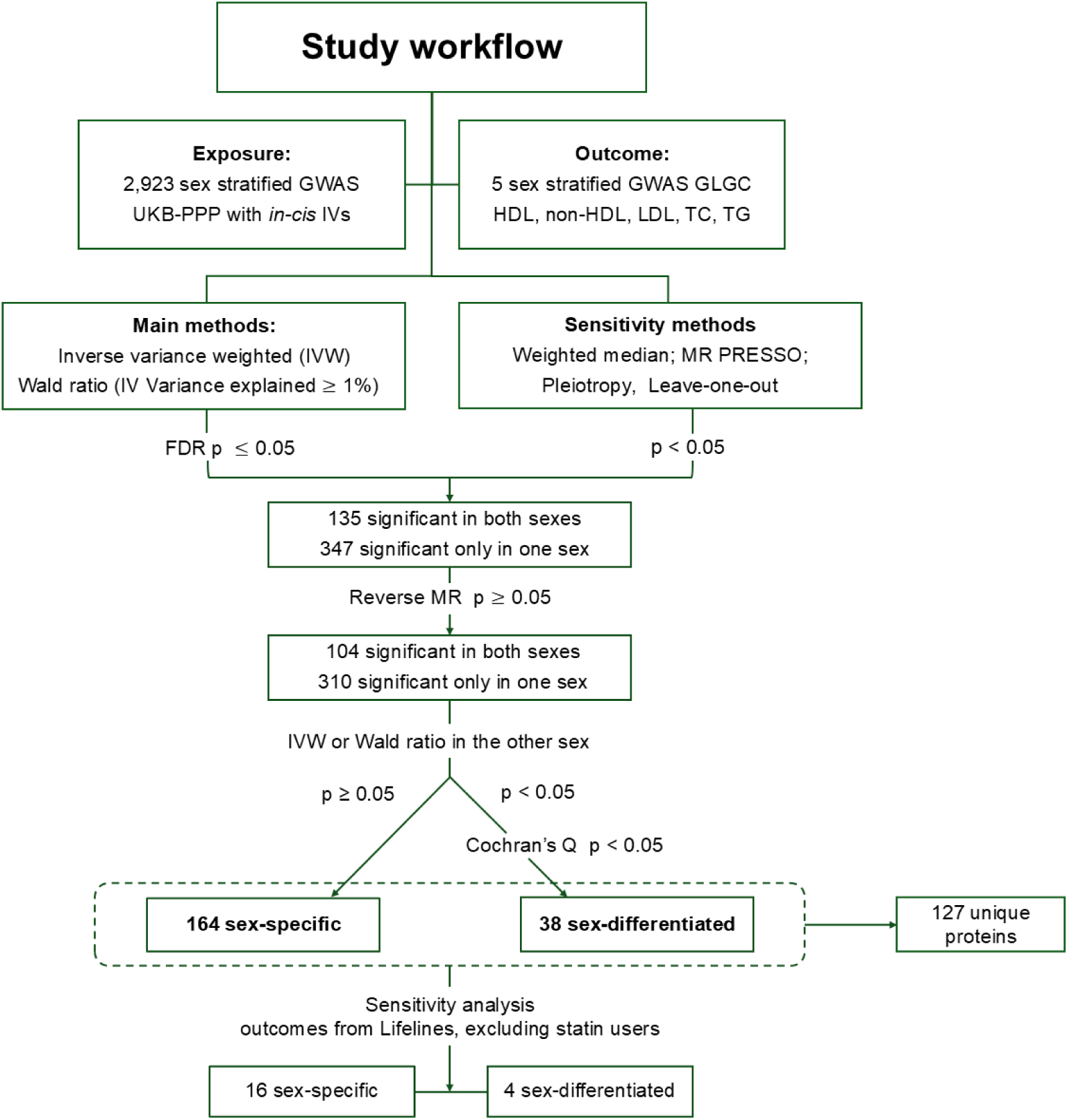
Flow chart of the different data sources and methods used in this study. The figure provides a schematic representation of the methods, filtering steps and results obtained. Abbreviations: GWAS, genome-wide association studies; UKB-PPP, UK Biobank Pharma Proteomic Project; HDL, high-density lipoprotein cholesterol; non-HDL, non-high-density lipoprotein cholesterol; LDL, low-density lipoprotein cholesterol; TC, total cholesterol; TG, triglyceride; GLGC, Global Lipid Genetic Consortium; IVs, instrument variables.

## Results

### Mendelian randomization analyses

A total of 1,882 and 1,910 proteins were associated with at least one cis genomic variant in males and females, respectively, in the UKB-PPP dataset and were therefore selected as exposures for two-sample MR analyses. MR was carried out using all potential IVs acting only in cis (Supplementary Table 1).

After removing causal relationships driven by just one single IV (Supplementary Table 2), we identified a total of 347 non-pleiotropic (p-value ≤ 0.05 in MR Egger pleiotropy test) formally significant causal relationships in only one sex (FDR≤ 0.05 in IVW and Wald ratio, nominal p-value in weighted median and MR-PRESSO), while in the other sex the evidence of an effect was null or not conclusive (Supplementary Table 3). We further removed relationships showing evidence of reverse causality (Supplementary Table 4) and kept a total of 310 non pleiotropic links; among them, 164 met our strict criteria for being defined sex-specific (IVW p-value or Wald ratio ≥0.05 in the other sex) (82 male-only and 82 female-only) (Supplementary Table 5 and Figure 1).

Scatter plots and leave-one-out plots performed across all MR methods tested in our study are available at https://github.com/Sanna-s-LAB/Mendelian-randomization-Project/tree/main/Zanetti_et_al_2025/Supplementary%20material.

We evaluated effect size differences between males and females for the 146 causal relationships formally significant only in one sex but not acting as sex-specific (310-164), and additional 104 relationships that were formally significant in both sexes. We used the Cochran’s Q test on the estimated causal effects from the main MR methods: IVW and Wald ratio. We identified 38 relationships with significant differences in causal estimates (Cochran’s Q p<0.05) (Supplementary Table 5). For example, the three proteins with the most pronounced sex differentiated causal estimates were: Neurocan core protein (NCAN), on LDL, non-HDL, TC, and TG; Cadherin EGF LAG seven-pass G-type receptor 2 (CELSR2) on non-HDL; and Killer cell immunoglobulin-like receptor 3DL1 (KIR3DL1) on TG (full results in Supplementary Table 5). With the main aim of investigating differences in causal relationships between sexes, in Figure 2 we highlighted the 164 (82 females-only, 82 males-only) significant sex-specific protein-lipid links detected by the MR analyses and in Figure 3 we showed the 38 significant relationships detected by the Cochran’s Q test. Overall, we observed that the direction of the causal estimates was generally consistent between females and males, and the sex differences laid in the magnitude. However, for a total of 41 relationships, we detected opposite estimate between the sexes (see “yes” in the “opposite_estimate” column in Supplementary Table 5)— for example, the protein NCAN showed a negative causal estimate for HDL in males and a positive estimate in females, whereas Allograft Inflammatory Factor 1 (AIF1) showed negative estimates for non-HDL and LDL in females and positive causal estimates in males.

**Figure 2.**
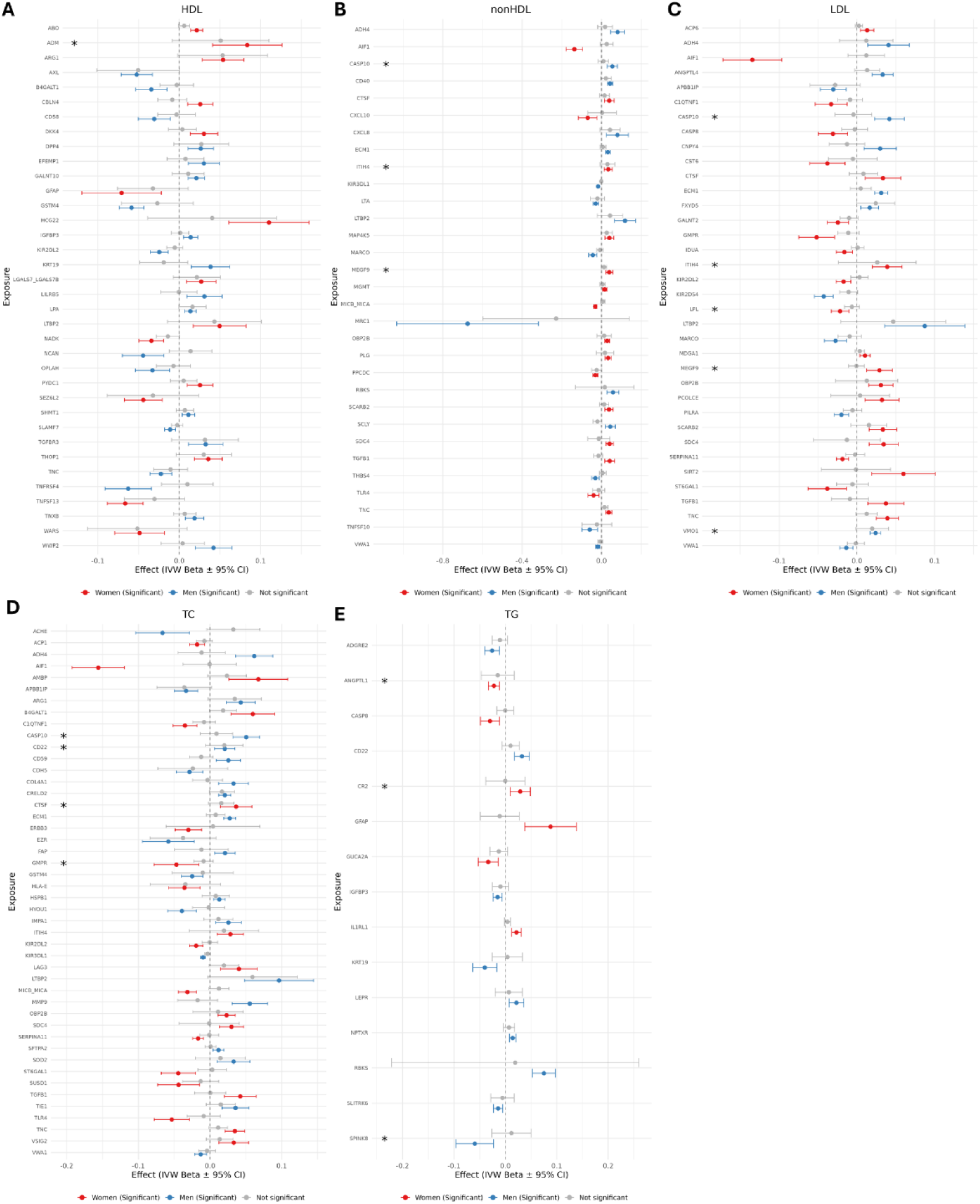
Forest plot of the proteins showing sex-specific causal effects. The figure provides a graphical representation of causal estimates according to IVW method and corresponding confidence intervals of protein effect on lipid levels for all the significant causal links detected. A, HDL, high-density lipoprotein cholesterol; B, non-HDL, non-high-density lipoprotein cholesterol; C, LDL, low-density lipoprotein cholesterol; D, TC, total cholesterol; E, TG, triglyceride * Indicates significant replication in the Lifelines cohort excluding individual under lower lipid medication treatment (statins). Abbreviations: IVW, inverse variance weighted method.

**Figure 3.**
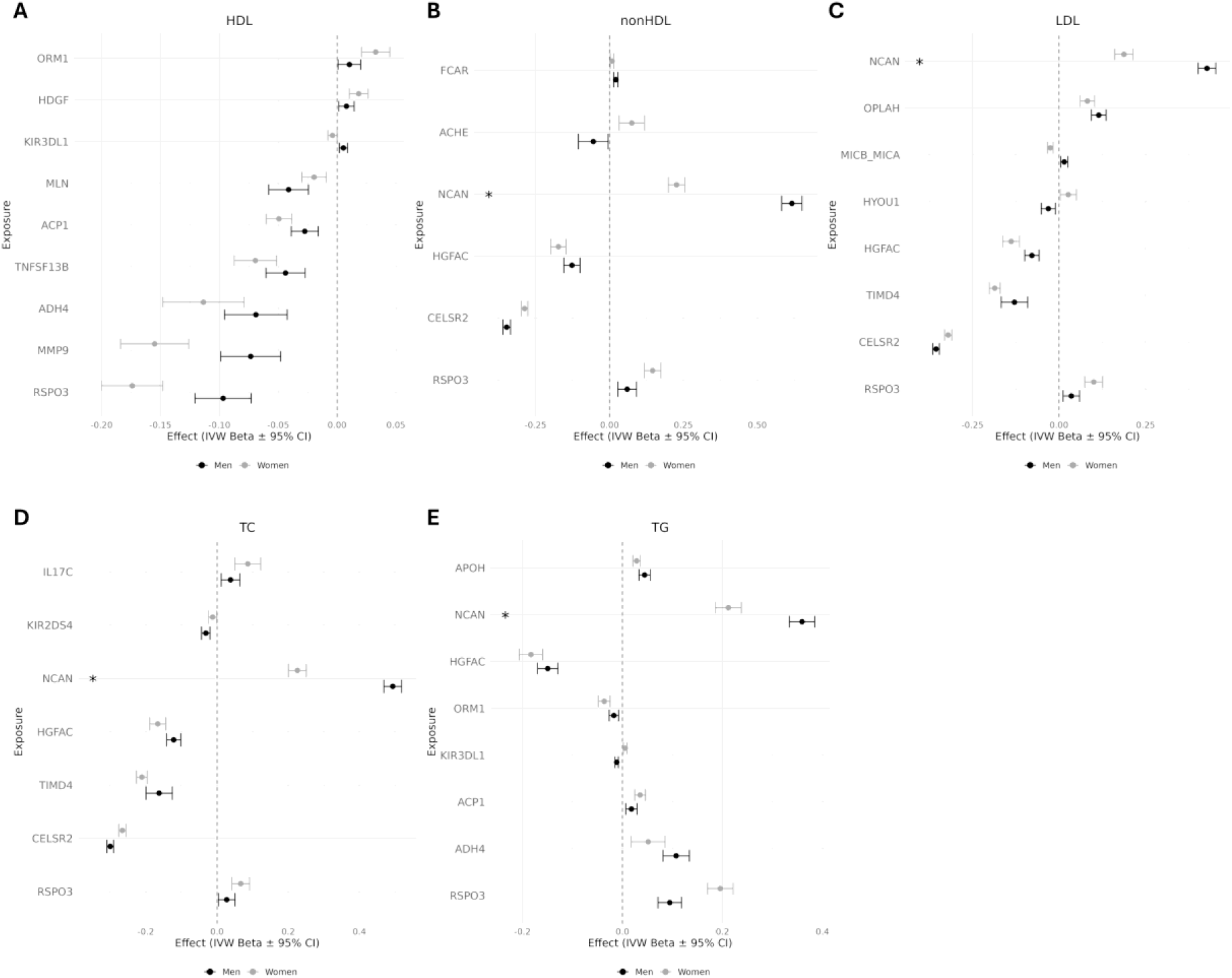
Forest plot of the proteins showing sex differentiated effects. The figure provides a graphical representation of causal estimates according to IVW method and corresponding confidence intervals of protein effect on lipid levels for all the proteins showing significant differences in causal estimate detected by Cochran’s Q test. A, HDL, high-density lipoprotein cholesterol; B, non-HDL, non-high-density lipoprotein cholesterol; C, LDL, low-density lipoprotein cholesterol; D, TC, total cholesterol; E, TG, triglyceride * Indicates significant replication in the Lifelines cohort excluding individual under lower lipid medication treatment (statins). Abbreviations: IVW, inverse variance weighted method

Among the identified proteins, we noted known players in lipid metabolism like the lipoprotein lipase (LPL) associated with increased levels of LDL only in women or the Apolipoprotein(a) (LPA) associated with increased levels of HDL and TG only in males. In addition, we detected novel important relationships with lipids, like the Leptin Receptor (LEPR), a key player in the regulation of energy balance and body weight control ^23^ and the Matrix metalloproteinase-9 (MMP9), implicated in atherosclerotic plaque rupture ^24^, showing in our study an higher causal effect on TG and on TC and LDL, respectively only in males.

### Replication in the Lifelines cohort

We performed a GWAS of lipid levels in the Lifelines Study Cohort to replicate our MR results and to check if usage of statins (lipid-lowering medications) could have introduced bias in our causal estimates. In this cohort, we tested for replication the 164 sex-specific and the 38 sex-differentiated relationships. We declared replication of sex-specific when the causal relationship was considered sex-specific according to our definition in the same sex-group and with the same direction as in the main analysis. We declared replication of sex-differentiated when the Cochran’s Q test p was <0.05 and the direction of single causal estimates were the same as in the main analysis.

When using the sex-specific GWASs derived in the full Lifelines Study cohort (N ≃ 38,554 in females and N ≃ 25,647 in males) we successfully replicated a total of 20 relationships, 13 in females and 7 in males. All the 20 sex-specific relationships detected except 4 also replicated when using the GWAS derived excluding statin users (N ≃ 36,982 in females and N ≃ 23,895) (see “yes” in the column Consistency_GLGC_Lifelines_sexspecific in Supplementary Table 6). The four relationships that did not show replication after removing statin users involve the following three proteins: Syndecan-4 (SDC4), FXYD domain-containing ion transport regulator 5 (FXYD5), and Cathepsin F (CTSF). A direct link between these six proteins and statins was not reported previously. A total of 4 sex-differentiated relationships replicated in the Lifelines cohort in the full dataset ad excluding individuals using statins (see “yes” in the column Consistency_QCochran_Lifelines_sexspecific in Supplementary Table 6).

### Characterization of proteins selected

We identified a total of 127 unique proteins from the previous analyses, specifically 116 acting sex-specific and 21 in a sex-differentiated manner; 10 of them behaved as sex-specific or sex-differentiated depending on the outcome. A full list of the 127 significant proteins together with their function is shown in Supplementary Table 7. Several of these proteins are directly linked to lipid metabolism, such as LPL ^25^, LPA ^26^, and LEPR ^27^. Others, including C-X-C motif chemokine ligand 8 (CXCL8) ^28^, are associated with inflammation—a key contributor to metabolic syndrome. Additional notable proteins include insulin-like growth factor binding protein 3 (IGFBP3), which has been linked to T2D ^29^, and MMP9, which is implicated in atherosclerosis ^30^.

Of note, in all the outcomes, except non-HDL and LDL, males showed the highest number of sex-specific proteins detected (Figure 4A). Moreover, most proteins within each sex were outcome-specific and not shared across the five lipid-related outcomes analyzed (Figure 4B). In females, for example, 39 proteins were uniquely associated with a single lipid trait, with the highest number (14 proteins) linked exclusively to HDL, while only 18 proteins were associated with more than one lipid trait.

**Figure 4.**
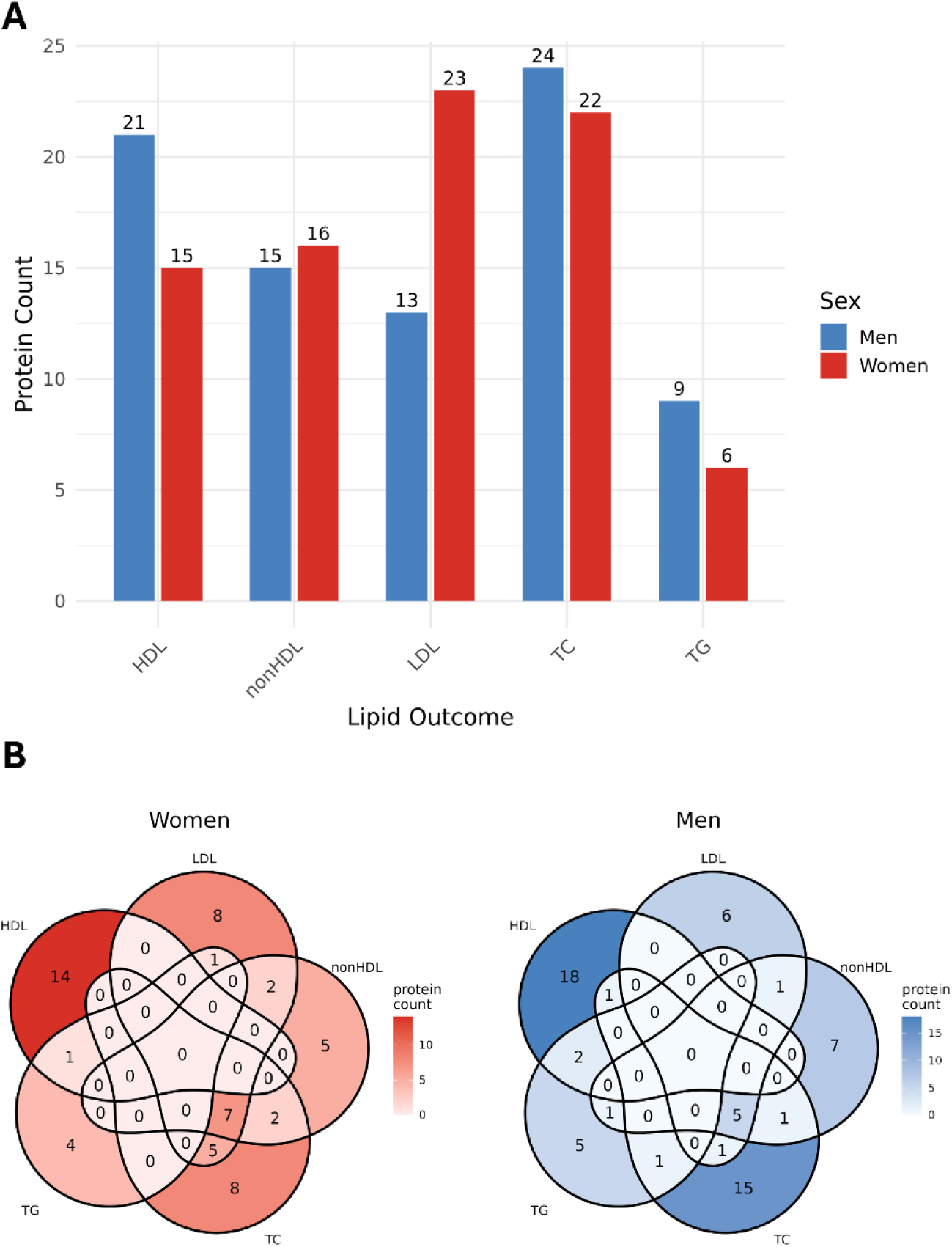
Sex-specific impact of causal relationships. A. The barplot provides a count of proteins selected in each outcome divided by sex. B. Venn diagrams showing the number of proteins shared or specific to lipid traits in males and females. For this plot in both panels, only sex-specific relationships are considered. Abbreviations: HDL, high-density lipoprotein cholesterol; non-HDL, non-high-density lipoprotein cholesterol; LDL, low-density lipoprotein cholesterol; TC, total cholesterol; TG, triglyceride.

Notably, none of the sex-specific proteins was causally linked with all five lipid traits together, in either sex. When outcomes were not considered, only 5 out of the 116 proteins were causally linked to both females and males, underscoring that the sex-specific relationships detected exist not only for the exposure–outcome pairs but also for individual proteins.

## Discussion

We studied the sex-specific causal relationships between 2,923 circulating proteins measured in the UKB-PPP, and lipid metabolism, specifically with HDL, non-HDL, LDL, TC and TG using GWASs from the Global Lipids Genetics Consortium.

Our main findings are the following: (1) we identified a total of 82 and 82 robust causal protein-lipids causal links that are specific to females and males; (2) a total of 38 protein– lipids links showed sex-differentiated causal effects; (3) the identified links corresponding to 127 unique proteins, specifically 116 sex-specific, 21 acting in sex-differentiated manner on lipid levels and 10 acting as sex-specific or sex-differentiated depending on the outcome. Most sex-specific proteins within each sex were outcome-specific and not shared across the five lipid-related outcomes analyzed. We also observed an higher proportion of sex-specific relationships in males for all traits (except for LDL and non-HDL), suggesting that, in females, regulation of lipids is more strongly influenced by non-genetic factors, for example sex hormones fluctuations during life stage. Of note, the difference in discovery is unlikely to be due to differences in statistical power, since the sample size of females-only GWASs was overall similar than the males-GWAS for both proteins and lipid GWASs.

### Comparison with prior literature

Among the proteins identified in our study, some are involved in various physiological processes, and their levels or activity are differently expressed in males versus females. MMP9 for example, emerged as a key candidate, being selected as sex-specific for TC. Specifically, we highlighted a causal relationship between increased levels of this protein and increased levels of TC in males (Figure 2D). A previous study highlighted that male rat aortic smooth muscle cells exhibit significantly higher MMP9 gene expression, protein levels, and enzymatic activity compared to female^31^. However, to our knowledge no published evidence exist on the opposite causal effect of MMP9 on TC in males and females.

Another protein worth mentioning, is the Angiopoietin-like 4 (ANGPTL4), a multifunctional glycoprotein that plays key roles in lipid metabolism, vascular biology, and inflammation^32^. It has been previously seen in mice that ANGPTL4 expression is higher in males than females in both liver and adipose tissue, and this difference is functionally relevant—male mice show a greater shift in plasma triglycerides and cholesterol when ANGPTL4 is genetically altered^33^. In line with this, our study highlighted a causal link of increased levels of ANGPTL4 on increased levels of LDL only in males (Figure 2C).

Lipid levels have recently emerged as independent factors contributing to the onset of heart failure with preserved ejection fraction^34^. A previous study highlighted that in heart failure with preserved ejection fraction, females exhibited higher circulating LPL and IGFBP-3 levels than males^35^. In our study we detected a causal relationship of increased IGFBP-3 levels with increased level of HDL (Figure 2A) and decreased levels of TG in males only (Figure 2E) and a causal sex-specific link of LPL and lower LDL in females which replicate also in the Lifelines cohort (Figure 2C). Other previous evidence suggested that LPL activity shows a clear sex difference with fundamental differences in the regulation of triglyceride uptake between males and females in adipose regions. Females show stronger associations between adipocyte size and LPL activity in specific fat depots, whereas males show different regional patterns^36^. Therefore, our results suggest that a differential molecular effect of these proteins could be involved in sex dimorphism of heart failure with preserved ejection fraction. In addition, we found evidence of sex-specific causal relationship of the Toll-Like Receptor 4 (TLR4) and decreased levels of TC (Figure 2D) and non-HDL (Figure 2B) only in females. Emerging evidence indicates that TLR4 signaling is modulated by sex hormones and exhibits sex-specific regulatory patterns with implications for cardiovascular aging and immune function ^37^. Toll-like receptors (TLRs) comprise a family of pattern recognition receptors that, upon activation, initiate pro-inflammatory signaling cascades. These receptors are broadly expressed within the cardiovascular system, where accumulating data implicates them in the sexual dimorphism observed in cardiovascular pathophysiology. Notably, sex-specific differences in TLR activity persist with advancing age, suggesting a potential paradigm shift in understanding the mechanisms that underlie sex differences in cardiovascular aging^37^.

### Strengths and Limitations

Our study is the largest and most comprehensive study of sex-specific causal associations of protein biomarkers with lipid metabolism to date. Other strengths of our study include the large sample size, the most recent and powerful GWAS summary statistics used as exposures and outcomes, the range of sensitivity analyses increasing robustness of our findings and the replication of the results in a different cohort. We applied robust methods with special assumptions about the behavior of pleiotropic variants, such as Egger intercept ^38^, which assumes pleiotropic effects are uncorrelated with the genetic associations with the risk factor, and the MR-PRESSO ^19^, which excludes outlier variants as being potentially pleiotropic. In addition, we performed reverse MR to exclude any possible bias due to reverse causation or confounding, thereby strengthening the validity of our results.

We also acknowledge the study limitations. First, we were limited to using pQTLs currently available in the UKB-PPP, and thus missed proteins not measured in this cohort. Second, the participants included in the genetic analyses were of European ancestry and the age range of UKB-PPP participants is very narrow (49-60 years old). Hence, our results may not be generalizable to other ethnic groups with significantly different ages and different lipid metabolism profiles. Finally, we performed replication using a different data set as outcome: the Lifelines cohort (with and without individuals under lower-lipid medication treatment). While this provides already some evidence of results being genuine, a formal replication would require to also change the exposure cohort (UKB-PPP).

### Conclusions

Our work highlighted several proteins with sex-specific and sex-differentiated causal impact on lipid levels, results that could be obtained only with the availability of sex-specific GWASs. The discovery of these sex-differences can provide important etiological insights into the management of lipid metabolism. Such knowledge may in turn improve the predictive use of sex-specific pQTLs and point to new therapeutic gender specific strategies to prevent CVD. Finally, we enhance the need to perform sex-stratified pQTL mapping in all future GWAS of any complex trait and disease as route to derive relevant insights into potential genetic drivers of sex dimorphism not only in lipid metabolism.

## Ethics approval and consent to participate

For the main analyses of this study only summarized non-individual data was used, and thus ethical approval was not needed. For replication purposes we used individual level data from the Lifelines Cohort Study, under project number OV23_00790. The Lifelines study was approved by the medical ethical committee of the University Medical Center Groningen (METc number: 2017/152).

## Consent for publication

Not applicable.

## Availability of data and materials

Sex-stratified GWAS summary statistics for all proteomic measurements available through UKBB-PPP are available at omicscience.org. Further information can be found at Koprulu et al. (2025)^8^. GWASs data from the GLGC consortium were downloaded from https://csg.sph.umich.edu/willer/public/glgc-lipids2021/results/sex_and_ancestry_specific_summary_stats/. The scripts used for all the analyses to reproduce the main results, as well as additional figures are publicly available in the GitHub repository: https://github.com/Sanna-s-LAB/Mendelian-randomization-Project/tree/main/Zanetti_et_al_2025.

Conversely, the Lifelines Cohort Study data that was used for replication is not available for direct download and can be requested, for a fee, by filling the application form at https://www.lifelines.nl/researcher/how-to-apply/apply-here. Lifelines will not charge an access fee for controlled access to the full dataset used in the manuscript, for the specific purpose of replication of the results presented in this Article or for further assessment by the reviewers, for a period of three months. Researchers interested in such a replication study or review assessment can contact Lifelines at.

## Declaration of interests

The authors declare no competing interests.

## Data Availability

Sex-stratified GWAS summary statistics for all proteomic measurements available through UKBB-PPP are available at omicscience.org. Further information can be found at Koprulu et al. (2025). GWASs data from the GLGC consortium were downloaded from https://csg.sph.umich.edu/willer/public/glgc-lipids2021/results/sex_and_ancestry_specific_summary_stats/. The scripts used for all the analyses to reproduce the main results, as well as additional figures are publicly available in the GitHub repository: https://github.com/Sanna-s-LAB/Mendelian-randomization-Project/tree/main/Zanetti_et_al_2025. Conversely, the Lifelines Cohort Study data that was used for replication is not available for direct download and can be requested, for a fee, by filling the application form at https://www.lifelines.nl/researcher/how-to-apply/apply-here. Lifelines will not charge an access fee for controlled access to the full dataset used in the manuscript, for the specific purpose of replication of the results presented in this Article or for further assessment by the reviewers, for a period of three months. Researchers interested in such a replication study or review assessment can contact Lifelines at.

https://github.com/Sanna-s-LAB/Mendelian-randomization-Project/tree/main/Zanetti_et_al_2025/Supplementary%20material

## Acknowledgements

This study was supported by Marie-Curie Fellowship to D.Za., acronym ‘Sex Dimorphism’ (GA n. 101066678) and by grants DSB AD006.371 “InvAt” FOE 2022 and grants ERC Stg n. 101075624 to S.S. The Lifelines initiative has been made possible by subsidy from the Dutch Ministry of Health, Welfare and Sport, the Dutch Ministry of Economie Affairs, the University Medical Center Groningen (UMCG), Groningen University and the Provinces in the North of the Netherlands (Drenthe, Friesland, Groningen).

We thank Davide Murrau for continuous support and management of our IT infrastructure. The authors wish to acknowledge the services of the Lifelines Cohort Study and of the UK Biobank Pharma Proteomics Project, the contributing research centers delivering data to Lifelines, and all the study participants. Lifelines access was granted via project number OV23_00790.

## Author’s contributions

DZa and SS designed the study and provided funding;

DZa and FG performed GWAS data QC and Mendelian Randomization analyses;

DZa and FG worked on data visualization;

MK analyzed UKBB data;

DZh analyzed the Lifelines cohort;

DZa and SS wrote the original manuscript draft;

MK DZh and FC supported data interpretation and provided critical revisions to the manuscript.

## Supplemental Material

### Supplementary Tables

**Supplementary Table 1.** Mendelian randomization results for all proteins in females, in males and in the full dataset.

The table reports the results for all proteins analyzed in the main MR analysis. The column SignificantBysex highlighted the significant results based on sex: 1, significant results in females; 2, significant results in males; 3, significant results in both sexes; 4, significant results showing a variance explained < 0.01 in one of the two sexes; 5, no significant results.

Abbreviations: HDL, high-density lipoprotein cholesterol; non-HDL, non-high-density lipoprotein cholesterol; LDL, low-density lipoprotein cholesterol; TC, total cholesterol; TG, triglyceride; nsnp, number of single nucleotide polymorphisms; b, causal effect estimate; se, standard error, pval, p-value, ple, pleiotropy; fdr, false discovery rate; het, heterozygosity; IV, instrument variable; exp_var, variance explained.

**Supplementary Table 2.** Leave-one-out results of the significant causal relationships.

The table lists leave-one-out results for all protein-lipid relationships showing FDR <0.05 and p-value >0.05 in the reverse MR analyses. The column “driving_SNP” indicated results that failed the leave-on-out test (yes), and the ones that failed the leave-one-out test but were excluded by MR-PRESSO (excluded_by_presso).

Abbreviations: HDL, high-density lipoprotein cholesterol; non-HDL, non-high-density lipoprotein cholesterol; LDL, low-density lipoprotein cholesterol; TC, total cholesterol; TG, triglyceride; snp, single nucleotide polymorphisms; b, causal effect estimate; se, standard error, pl, p-value; W, females, M, males.

**Supplementary Table 3.** Mendelian randomization significant results in females,males and in both sexes.

The column “SignificantBysex” highlighted the significant results based on sex in the three different analyses: 1, significant results in females; 2, significant results in males; 3, significant results in both sexes; 4, significant results showing a variance explained < 0.01 in one of the two sexes; 5, no significant results.

Abbreviations: HDL, high-density lipoprotein cholesterol; non-HDL, non-high-density lipoprotein cholesterol; LDL, low-density lipoprotein cholesterol; TC, total cholesterol; TG, triglyceride; nsnp, number of single nucleotide polymorphisms; b, causal effect estimate; se, standard error, pval, p-value, ple, pleiotropy; fdr, false discovery rate; exp_var, variance explained.

**Supplementary Table 4.** Reverse Mendelian randomization results of the significant causal links from Supplementary Table 1.

The table reports the results for the reverse MR analyses, e.g. using the lipids GWAS as exposure and the protein GWAS as outcomes. The column SignificantBysex highlighted the significant results based on sex: 1, significant results in females; 2, significant results in males; 3_1, significant results in both sexes in the main cist+trans analyses and only significant in females in the bidirectional analyses; 3_2, significant results in both sexes in the main cist+trans analyses and only significant in males in the bidirectional analyses, 3_3, significant results in both sexes in the main cist+trans and in the bidirectional analyses; 4_1, significant results in the main cis+trans analyses showing a variance explained < 0.01 in one of the two sexes and only significant in females in the bidirectional analyses; 5, no significant results.

Abbreviations: HDL, high-density lipoprotein cholesterol; non-HDL, non-high-density lipoprotein cholesterol; LDL, low-density lipoprotein cholesterol; TC, total cholesterol; TG, triglyceride; nsnp, number of single nucleotide polymorphisms; b, causal effect estimate; se, standard error, pval, p-value, ple, pleiotropy; fdr, false discovery rate; het, heterozygosity; IV, instrument variable; exp_var, variance explained.

**Supplementary Table 5.** Cochran’s Q test results for the significant MR causal relationships.

The column “SignificantBysex” highlighted the significant results based on sex in the three different analyses: 1, significant results in females; 2, significant results in males; 3, significant results in both sexes; 4, significant results showing a variance explained < 0.01 in one of the two sexes; 5, no significant results. The table shows the absolute difference of the betas between males and females (Diff_Beta) and the Cochran’s Q test results applied to the weighted betas.

Abbreviations: HDL, high-density lipoprotein cholesterol; non-HDL, non-high-density lipoprotein cholesterol; LDL, low-density lipoprotein cholesterol; TC, total cholesterol; TG, triglyceride; nsnp, number of single nucleotide polymorphisms; b, causal effect estimate; se, standard error, pval, p-value, ple, pleiotropy; fdr, false discovery rate; exp_var, variance explained.

**Supplementary Table 6.** Mendelian randomization results using proteins from the UK Biobank Pharma Proteomic Project (UKB-PPP) as exposures and lipid traits in the Lifelines cohort as outcome.

A “yes” in the column “Consistency_GLGC_” indicated a replication of the significant result considering the same sex-specific result as in the main analysis, and same direction of causal effect in the whole Lifelines cohort (_all), additionally excluding individuals under lower-lipid medication treatment (_nostatins) and a replication in both Lifelines datasets, the entire cohort and excluding individuals with statins (_Lifelines).

Abbreviations: HDL, high-density lipoprotein cholesterol; non-HDL, non-high-density lipoprotein cholesterol; LDL, low-density lipoprotein cholesterol; TC, total cholesterol; TG, triglyceride; nsnp, number of single nucleotide polymorphisms; b, causal effect estimate; se, standard error, pval, p-value, ple, pleiotropy; fdr, false discovery rate; het, heterozygosity; IV, instrument variable; exp_var, variance explained; GLGC, Global Lipid Genetic Consortium.

**Supplementary Table 7.** Sex specific and sex differentiated proteins selected in the different analyses with their function.

### Supplementary Figures

**Supplementary Figures 1.** Scatter plots performed across all MR methods tested in our study and the leave-one-out sensitivity analysis for the MR carried out. Each black point in the forest plot represents the MR analysis (using the inverse variance weighting method) excluding that particular SNP. The overall analysis including all SNPs is also shown for comparison. **A**. In females. **B**. In males. **C**. In the full dataset.

**Supplementary Figures 2.** Scatter plots performed across all MR methods tested in our study and the leave-one-out sensitivity analysis in the reverse MR analyses. Each black point in the forest plot represents the MR analysis (using the inverse variance weighting method) excluding that particular SNP. The overall analysis including all SNPs is also shown for comparison. **A**. In females. **B**. In males.

**Supplementary Figures 3.** Scatter plots performed across all MR methods tested in our study and the leave-one-out sensitivity analysis in the Lifelines cohort. Each black point in the forest plot represents the MR analysis (using the inverse variance weighting method) excluding that particular SNP. The overall analysis including all SNPs is also shown for comparison. **A1**. Results obtained when using the females-only GWAS. **A2**. Results obtained when using the females-only GWAS but excluding subjects under lower-lipid medication treatment. **B1**. Results obtained when using the males-only GWAS. B**2**. Results obtained when using the males-only GWAS but excluding subjects under lower-lipid medication treatment.

## Notes

### Competing Interest Statement

The authors have declared no competing interest.

